# Biomimetic stimulation patterns drive natural artificial touch percepts using intracortical microstimulation in humans

**DOI:** 10.1101/2024.07.31.24311276

**Authors:** Taylor G. Hobbs, Charles M. Greenspon, Ceci Verbaarschot, Giacomo Valle, Michael Boninger, Sliman J. Bensmaia, Robert A. Gaunt

## Abstract

**Objective:** Intracortical microstimulation (ICMS) of human somatosensory cortex evokes tactile percepts that people describe as originating from their own body, but are not always described as feeling natural. It remains unclear whether stimulation parameters such as amplitude, frequency, and spatiotemporal patterns across electrodes can be chosen to increase the naturalness of these artificial tactile percepts.

**Approach:** In this study, we investigated whether biomimetic stimulation patterns – ICMS patterns that reproduce essential features of natural neural activity – increased the perceived naturalness of ICMS-evoked sensations compared to a non-biomimetic pattern in three people with cervical spinal cord injuries. All participants had electrode arrays implanted in their somatosensory cortices. Rather than qualitatively asking which pattern felt more natural, participants directly compared natural residual percepts, delivered by mechanical indentation on a sensate region of their hand, to artificial percepts evoked by ICMS and were asked whether linear non-biomimetic or biomimetic stimulation felt most like the mechanical indentation.

**Main Results:** We show that simple biomimetic ICMS, which modulated the stimulation amplitude on a single electrode, was perceived as being more like a mechanical indentation reference on 32% of the electrodes. We also tested an advanced biomimetic stimulation scheme that captured more of the spatiotemporal dynamics of cortical activity using co-modulated stimulation amplitudes and frequencies across four electrodes. Here, ICMS felt more like the mechanical reference for 75% of the electrode groups. Finally, biomimetic stimulation required less stimulus charge than their non-biomimetic counterparts.

**Significance:** We conclude that ICMS encoding schemes that mimic naturally occurring neural spatiotemporal activation patterns in somatosensory cortex feel more like an actual touch than non-biomimetic encoding schemes. This also suggests that using key elements of neuronal activity can be a useful conceptual guide to constrain the large stimulus parameter space when designing future stimulation strategies.

## 1. Introduction

Somatosensory feedback is essential for object manipulation [1–3], and without the sense of touch, many everyday movements become highly impaired [4]. Therefore, neuroprostheses that include artificial somatosensory feedback that is natural and intuitive will likely maximize embodiment and usability [5–8], and improve the performance of brain-controlled bionic limbs [9]. In the context of somatosensory neuroprosthetics, artificial tactile percepts that are indistinguishable from naturally occurring ones would require minimal learning or interpretation and would be more likely to support complex sensorimotor behaviors. For example, increasing the naturalness of artificial tactile percepts evoked by electrical stimulation of peripheral nerves in people with amputations led to improved control and embodiment of neuroprosthetic hands and legs [10–12].

Intracortical microstimulation (ICMS) of the somatosensory cortex in humans can restore somatosensory feedback after many types of injuries as it activates the cortex directly [13] and does not rely on a functional peripheral nervous system. Multiple labs have shown that ICMS can elicit artificial sensations that feel as if they come from an individual’s own hand (figure 1(a)) [13–17]. These sensations are stable over years [18], and while they can have natural qualities, such as pressure and touch [13,14,16,17] they can also evoke unnatural sensations such as tingling and buzzing [13,16,19]. Controlling the quality of these artificial sensations will likely be an important ability for future systems. At the simplest level, increasing the current amplitude increases the perceived intensity of the stimulus [13], and to some extent, the quality of artificial sensations can be changed by modulating the stimulation frequency and other features of the pulse trains [19–21]. In an ideal system, electrical stimulation should feel natural, but this is nontrivial to both create and measure. For practical purposes, a *natural* sensation can be defined as a sensation that feels like interactions with everyday objects.

**Figure 1.**
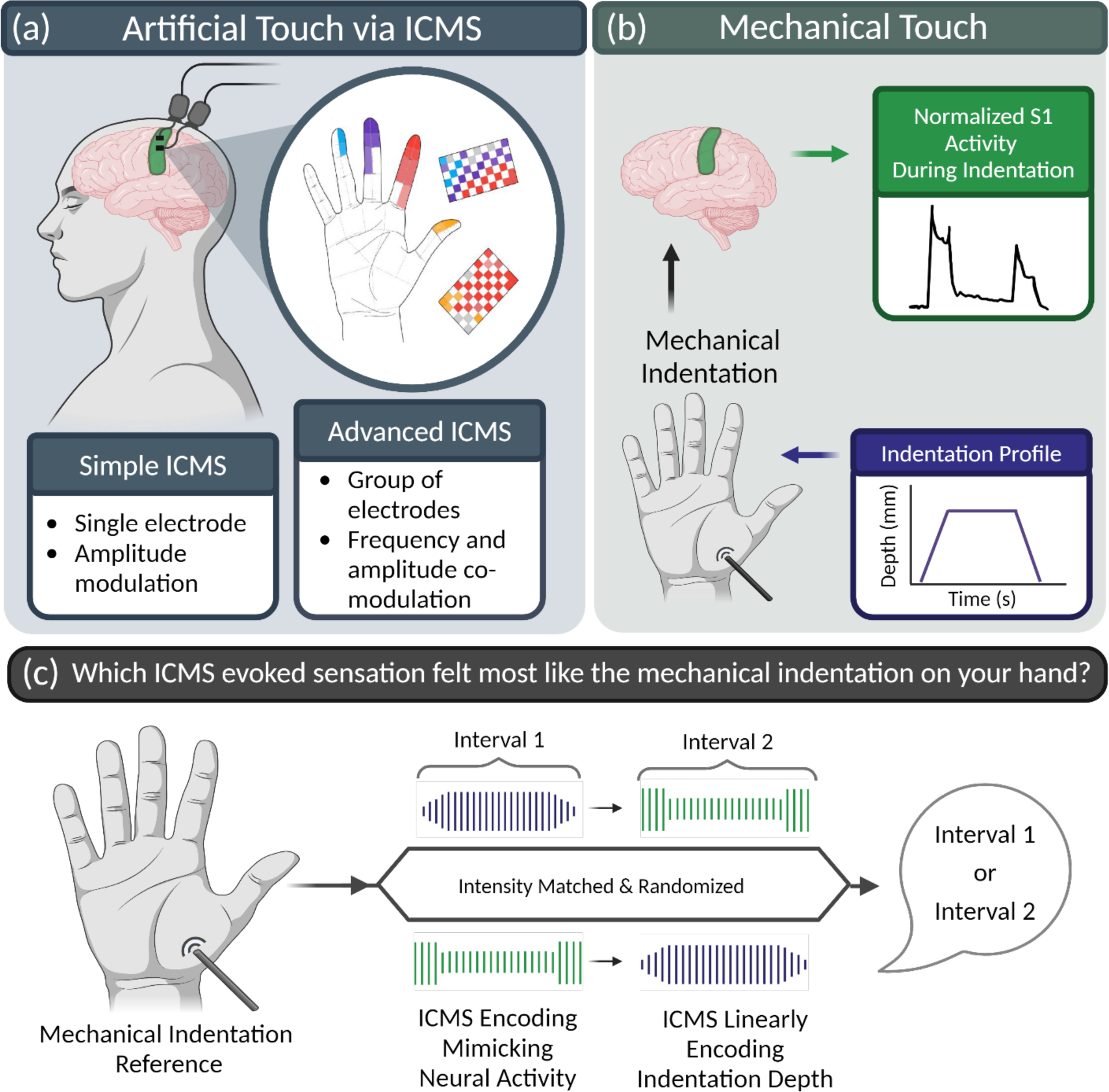
Graphical overview of experimental concept and testing paradigm. (a) ICMS delivered though two implanted microelectrode arrays in the somatosensory cortex (green region highlighted in cortex) elicits tactile sensations that are experienced as though they originate directly from the participants’ hands. Evoked sensations are topographically organized, corresponding to the electrode location on the array. Data from C1 (adapted from Greenspon, et al. 2024). Two encoding schemes were used for this study; simple and advanced ICMS. (b) The mechanical indentation profile (purple box) shows the periods of ramping up and down and is delivered to sensate regions on the participant’s palm. Neural activity in S1 during mechanical indentations on the hand is dominated by transient responses – spatiotemporally and in firing rate – at the stimulation onset and offset (green box, based upon Callier, et al., 2019). (c) Experimental design. Participants received a natural mechanical indentation on their hand to serve as the reference to evaluate naturalness. Two ICMS trains were then delivered encoding the stimulation parameters as a linear representation of the indentation depth (purple) or as a biomimetic representation of the cortical neural activity (green). Participants were asked to verbally report which artificial percept felt more like the reference sensation. Parts of this imaged were created with BioRender.com.

Attempting to use naturally evoked neural activity as the basis of ICMS patterns is often referred to as biomimetic stimulation and has already been shown to hold promise in rodents and humans. In rodents, modulating ICMS amplitude in the somatosensory cortex using a biomimetic encoding algorithm induced bursts of activity in neurons that parallel those observed during touch onset and offset [22]. This was also true if frequency and amplitude were co-modulated, but not if the stimulation frequency alone was modulated using a biomimetic encoding scheme. One motivation for co-modulating amplitude and frequency is that co-modulation enhances the difference in neural population activation between contact transients and sustained touch. Additionally, in humans, biomimetic ICMS in the somatosensory cortex results in improved tactile sensitivity, leading to better force feedback perception, especially with ICMS delivered through multiple electrodes [15]. Delivering stimulation across multiple electrodes explicitly mimics the spatial spread of activity seen in natural touch during contact transients [23]. Biomimetic ICMS has also been shown to have fewer detrimental effects related to stimulation artifact using closed loop decoders [24]. Finally, biomimetic stimulation patterns delivered to peripheral nerves can improve the naturalness of evoked tactile percepts [10,11].

Given these observed benefits, we sought to determine if biomimetic microstimulation in the somatosensory cortex would improve the perceived naturalness of a stimulation train compared to non-biomimetic ICMS. However, there are two critical challenges for developing ICMS trains that produce natural percepts. First, there is a vast multidimensional stimulation parameter space, with numerous combinations of amplitudes, frequencies, and temporal patterns across many electrodes. Second, quantifying naturalness itself presents a challenge, both conceptually and in terms of measurement accuracy, since determining the ‘naturalness’ of a sensation is not straightforward and is highly subjective.

A practical approach to exploring the stimulation parameter space is to guide decisions based on a theoretical framework. For an ICMS encoding scheme, this means defining a set of principles that determine how stimulation parameters are created given some input signal. A simple approach, linear encoding, relates the contact pressure measured from sensors on a prosthetic hand to the stimulation amplitude so that the perceived intensity of the stimulus increases as the contact pressure increases [9]. While this exploits the established linear relationship between amplitude and perceived intensity [13], it does not consider how populations of neurons in the somatosensory cortex naturally respond to normal touch. Indeed, both single neuron and population data from non-human primates shows that contact transients are far more powerful drivers of neural activity than sustained contact, both temporally and spatially (figure 1(b)) [23]. This transient-dominated neural activity is a key principle that provides information about object contact and interactions [1].

To address the problem of measuring naturalness, we developed a task that implicitly, but quantitatively assesses naturalness, and does not require a descriptive evaluation from the participants. In this task, we first provided mechanical stimulation on a sensate area on the participants’ skin as a reference for a natural sensation. We then delivered both linear and biomimetic ICMS patterns in a random order and asked participants to indicate which ICMS train most resembled the mechanical reference (figure 1(c)). If a participant reported that the interval with biomimetic ICMS felt more like the mechanical indentation, it would signify that biomimetic ICMS felt more natural.

Using these approaches, stimulation using individual electrodes revealed a modest preference towards biomimetic ICMS. When we extended the biomimetic encoding scheme to incorporate co-modulation of amplitude and frequency, as well as spatial patterning using multiple electrodes, participants chose biomimetic ICMS over linear ICMS as more closely resembling the mechanical touch far more frequently. Our findings indicate that biomimetic stimulation of human somatosensory cortex better replicates the perceived sensations that we experience in our daily lives.

## 2. Materials and methods

### 2.1 Participants

This study was conducted under an Investigational Device Exemption from the U.S. Food and Drug Administration as part of Clinical Trial NCT01894802 and was approved by Institutional Review Boards at the University of Pittsburgh and the University of Chicago. Informed consent was obtained for all participants before the start of the study. The purpose of this trial is to collect preliminary safety information and demonstrate that intracortical electrode arrays can be used by people with tetraplegia to both control external devices and generate tactile percepts from the paralyzed limbs. This manuscript presents the analysis of data that were collected during the participants’ involvement in the trial, but does not report clinical trial outcomes.

Participant P2 (male) presented with a C5 motor/C6 sensory ASIA B spinal cord injury (SCI) that occurred 10 years prior to implantation. He had been implanted for approximately eight years at the time of this study. No sensation remains in the ulnar region of the hand from digits 3-5 on both the palmar and volar surfaces but diminished light touch and deep sensation are preserved on the radial side (digits 1-2) and the thenar eminence of the palm (0.16 g with monofilament testing) [25,26].

Participant P3 (male) presented with a C6 ASIA B SCI that occurred 12 years prior to implantation. He had been implanted for approximately three years at the time of this study. No sensation remains in the ulnar region of the hand on both the palmar and volar surfaces but diminished light touch and deep sensation are preserved on the radial side with normal sensory thresholds at the thenar eminence (0.07 g with monofilament testing) [25,26].

Participant C1 (male) presented with a C4-level ASIA D SCI that occurred 35 years prior to implantation. He had been implanted for approximately three years at the time of this study. Monofilament testing revealed spared deep sensation but diminished light touch in the right hand and as well as normal sensory thresholds at the thenar eminence (0.04 g with monofilament testing) [25,26].

### 2.2 Cortical implants

Four microelectrode arrays (Blackrock Neurotech, Salt Lake City, UT, USA) were implanted in the left hemisphere of each of the three participants [13,27]. Two of the arrays were placed in Brodmann’s area 1 of the somatosensory cortex. These two arrays were 2.4 mm × 4 mm, with 60 electrodes arranged in a 6 × 10 grid. Each electrode shank was 1.5 mm long and coated with sputtered iridium oxide film [28]. The electrodes were wired in a checkerboard pattern on the array, such that ICMS could be delivered through 32 electrodes. The other two 10 × 10 microelectrode arrays were placed in the hand and arm areas of motor cortex. These electrode arrays were not used in this study. Two percutaneous connectors (Blackrock Neurotech, Salt Lake City, UT, USA) were mounted on the skull, each connected to both a motor array and a sensory array. The placement of the four arrays was based on imaging data from fMRI or magnetoencephalography experiments in which participants attempted to move their hand and arm. Final surgical placement was constrained by cortical topography and surface vascular anatomy [27,29].

### 2.3 Experimental Protocol

To quantitively measure how stimulation encoding schemes affected the perceived naturalness of artificial tactile sensations, without relying on open-ended verbal reports, we designed a task that required the participants to identify whether a linear or biomimetic stimulation train was a better perceptual match to a mechanical stimulus delivered to a sensate region of their hand (figure 1(c)). At the start of each day, we matched the perceived intensity of the different ICMS trains (see below) for all electrodes that would be tested during that session. Participants then had their right hand comfortably held in place with Velcro straps to minimize hand movement during mechanical indentations. The starting position of the mechanical indenter was then adjusted such that the intensity of the mechanical indentation was approximately equal to the intensity of ICMS. The mechanical indentation location was marked with a pen to ensure the mechanical indenter did not move throughout the testing session on a given day. Intensity matching is an important part of the protocol as it ensures that differences in perception cannot be simply ascribed to the intensity of the stimulus.

Each trial contained a mechanical reference followed by two intervals of ICMS, one linear and one biomimetic (in random order). The start of each trial was indicated with a white cross presented on a monitor while the mechanical stimulus was indicated by a purple cross and the ICMS stimuli by a green cross. These cues were provided to assist with learning the task progression, but the participants were not required to maintain their gaze on the monitor. All stimuli were separated by a 3 s inter-stimulus-interval. This method ensured that participants had a consistent natural reference throughout the experiment. On each day, 20 to 60 trials were collected for an electrode or electrode group in blocks of four trials. Participants were asked to report which interval of stimulation felt most like the mechanical reference stimulus (figure 1(c)). Participants could request breaks between blocks if necessary and were allowed to repeat any trials they were unsure about at the end of each block.

For participants P1 and P2, each electrode or electrode group was tested on three days, with 20 trials per day. For participant C1, we tested four electrodes and five electrode groups on three days with 20 trials per day. We then used a Friedman’s test to check for differences across days, and after finding none (p = 0.63 for single electrodes; p = 0.76 for electrode groups), we tested an additional eight electrodes and three electrode groups using 60 trials in a single day.

### 2.4 Mechanical Indentation

The mechanical indentations were delivered as ramp-and-hold trapezoids. All indentations were 1 s in duration, 2 mm deep, had a ramp duration of 0.2 s (10 mm/s), and had a 0.6 s hold phase between ramps. To ensure that the intensity of the mechanical stimulus was comparable to the electrical stimulus, we either adjusted the specific location of the indenter on the skin surface to evoke a clear tactile percept, or increased the depth of skin pre-indentation such that the intensity of the mechanical stimulus was equivalent to the electrical stimulus [23,30]. The mechanical indenter was positioned above the thenar eminence as this area was sensate for all participants.

For participants P2 and P3, indentations were delivered with a Dual-Mode Lever Arm System model 300C-LR-I (Aurora Scientific, Aurora, Ontario, Canada), operating in a displacement control mode. The mechanical indentation tip diameter was 2 mm. For participant C1, indentations were delivered with a X-DMQ-AE direct drive linear stage (Zaber Technologies, Vancouver, British Columbia), and the mechanical tip diameter was 5 mm.

### 2.5 Intracortical Microstimulation

Intracortical microstimulation pulses were generated using a CereStim C96 multi-electrode stimulation system (Blackrock Neurotech, Salt Lake City, UT, USA). Each stimulation pulse was current-controlled and charge balanced, with a cathodal leading phase (200 μs), an interphase period (100 μs), and a half amplitude anodal phase (400 μs). All ICMS trains lasted 1 s, corresponding to the duration of the mechanical indentation.

#### 2.5.1 Simple Biomimetic and Linear Encoding

Simple biomimetic ICMS encoding (figure 2(a) top) used an amplitude modulation scheme to drive neural activity during mechanical indentations, with higher currents being delivered during indentation and release, and lower currents otherwise [23]. Controlling the stimulus current effectively modulates the volume of tissue activated [31–33]. These transient bursts of activity last as long as the mechanical on- and off-ramps [23] and thus were 0.2 s long. To ensure that the stimulus was perceived during the hold phase and that the transients activated substantially more tissue, we set the amplitude of the hold phase to 40 μA, which is well above the detection threshold [18], and set the transient amplitude to 80 μA. The stimulation frequency was set to 250 Hz to maximize the charge delivered during the transient phases.

**Figure 2.**
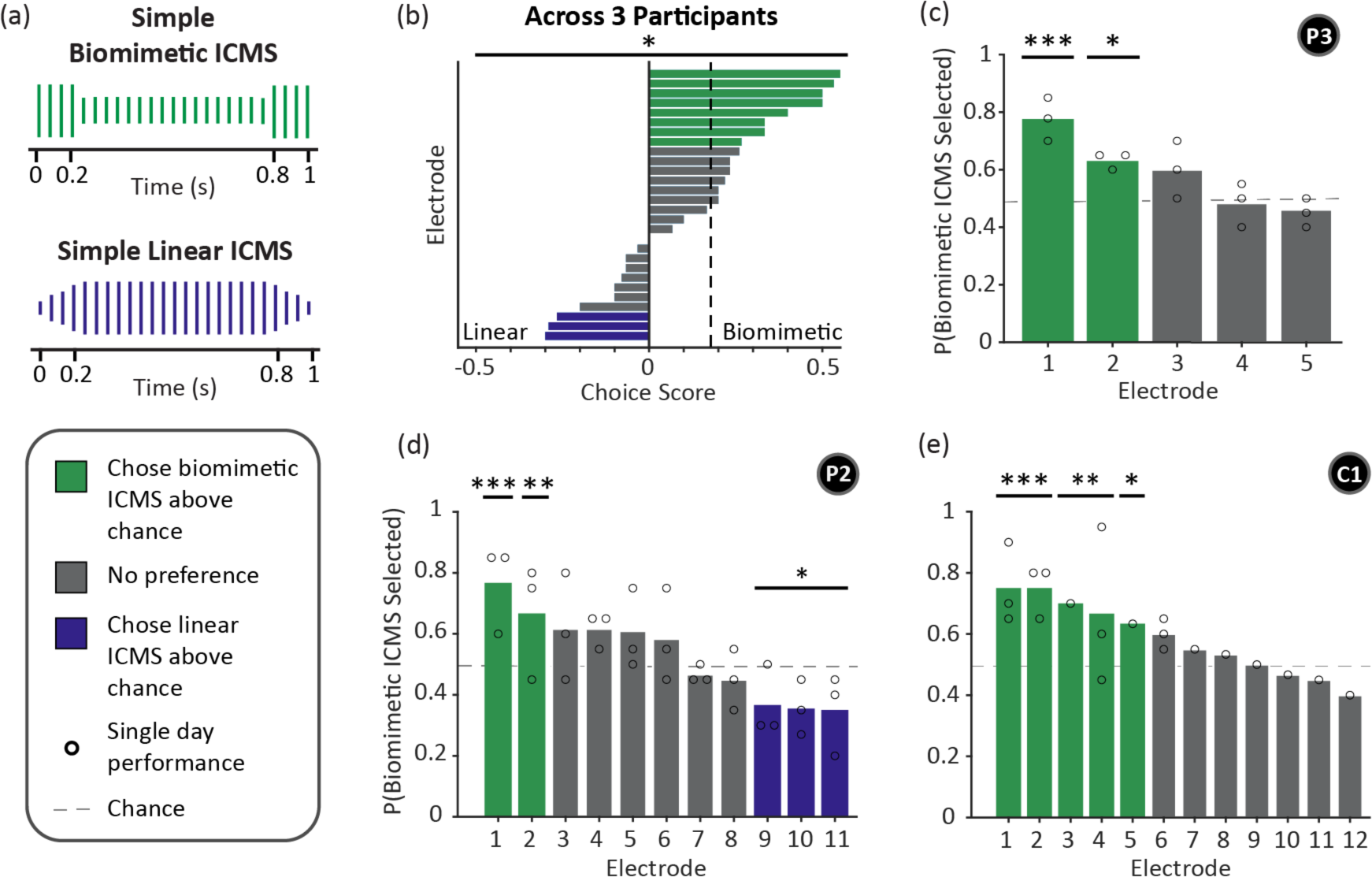
The effect of simple biomimetic and linear ICMS on the naturalness of ICMS-evoked tactile percepts. (a) All simple ICMS encoding schemes used a single electrode and modulated only the stimulus amplitude. Pulse amplitude is represented by the height of the colored lines. (Top) Simple biomimetic ICMS has bursts of stimulation to match the 0.2 s ramp durations of the mechanical indentation. (Bottom) Simple linear ICMS increased the stimulus amplitude in proportion to the depth profile of the mechanical indentation. (b) Choice scores for each electrode (see methods). Each bar indicates one electrode. The dashed line indicates the median choice score. A positive choice score indicates that biomimetic ICMS was a better perceptual match to the mechanical stimulus, while a negative choice score indicates that linear ICMS was a better perceptual match. A score of 0.5 indicates that biomimetic ICMS was selected on 75% of the trials. Bars are colored based on significance testing using a Chi-squared goodness of fit test (green = more biomimetic trials, gray = no difference, purple = more linear trials). A Wilcoxon signed rank test was used to determine if the choice scores differ from 0, indicating which ICMS encoding scheme produced more natural sensations (*p < 0.05). (c-e) Proportion of trials where biomimetic ICMS was selected as feeling more like the mechanical indentation reference across all testing days for each participant. Bar coloring is the same as above. Black circles show the proportion of trials for which biomimetic ICMS was chosen on a single day (Chi-squared goodness of fit test, ***p < 0.001, **p < 0.01, *p < 0.05). Participant identifiers included in black circles.

Simple linear ICMS encoding (figure 2(a) bottom) linearly modulated the stimulus amplitude based on the same ramp-and-hold profile used to control the mechanical indenter and represented the indentation depth. This is a plausible encoding method as perceived intensity scales with both ICMS current amplitude and indentation depth [9,13]. To ensure that the overall intensity of the linear and biomimetic ICMS trains were equivalent, we used a behavioral task in which the participant adjusted the maximum amplitude of the linear ICMS train (see below). The amplitude during the 0.2 s ramp phase was linearly interpolated between 2 μA and the maximum amplitude that participants selected during the intensity matching task. The frequency of simple linear ICMS was also set to 250 Hz.

#### 2.5.2 Advanced Biomimetic and Linear Encoding

In the simple biomimetic scheme, stimulation amplitude modulates the volume of tissue that is recruited by stimulation [31–33]. This captures only a single aspect of the natural cortical response to touch. Therefore, we also developed an advanced biomimetic ICMS (figure 4(b) left) scheme that co-modulated pulse amplitude and frequency across four electrodes to even better mimic two key features of neural activity during mechanical indentation; the increased firing rate of neurons and the increase in the spatial extent of responsive neurons during contact transients [23]. We chose to use four electrodes to deliver stimulation based on previous success with multi-electrode biomimetic ICMS [15]. The stimulation amplitude during the transient phases was again set to 80 μA. Between the onset and offset transients, only one electrode was used to deliver current at 40 μA. For these advanced biomimetic trains, the frequency during the transient phase was set to 200 Hz while during the hold phase the frequency was reduced to 100 Hz.

Advanced linear ICMS (figure 4(b) right) also incorporated frequency and amplitude co-modulation. Both the stimulus amplitude and frequency were scaled between their respective minimums and maximums (2 μA to the intensity matched amplitude, and 0 to 200 Hz, respectively) during the transient phases. All four electrodes delivered this stimulation pattern.

#### 2.5.3 Electrode Selection

For the simple stimulation trains, we chose electrodes that had detection thresholds below 40 μA to ensure that the biomimetic stimuli remained perceptible during the hold phase. In addition, we chose electrodes that were broadly distributed across both arrays in each participant to ensure that the results were not biased by potential changes in the local response to stimulation. For the advanced stimulus trains, we selected an additional three electrodes that were adjacent to each other, had detection thresholds below 40 μA, and evoked tactile percepts on similar areas of the hand.

### 2.6 Projected Fields

Projected fields are the location(s) on the hand where ICMS-evoked percepts are felt. To determine these projected fields, participants used a tablet to draw the spatial extent of the tactile percept on a digital representation of the hand in response to a 60 μA, 100 Hz, 1 s stimulus train. For multi-electrode projected fields, all four electrodes were stimulated simultaneously at 60 μA, 100 Hz, for 1 s. We averaged the projected field extents if data were collected multiple times during the duration of this experiment [29].

### 2.7 ICMS intensity matching

We sought to remove any confound associated with differences in perceptual intensity between the two ICMS encoding strategies. To accomplish this, participants matched the intensity of linear ICMS to biomimetic ICMS trains using a tablet interface at the start of each session. The biomimetic ICMS train was fixed, using the parameters described above, while the amplitude of the linear train could be adjusted with the slider. Participants were encouraged to adjust the slider until the intensity of the two ICMS trains was equivalent (approximating the method of adjustments [34]). This process was repeated both with the biomimetic train first and the linear train first to minimize order effects [35]. The final amplitude was averaged across the two repetitions. Participants reported no difficulty with the task using their residual motor capabilities.

### 2.8 Experimental analysis and statistics

Within a single session, we used a Chi-square goodness-of-fit test, with the prior distribution being equally proportionate, to determine whether biomimetic or linear ICMS trains were selected above chance (chance: 50%). We also used this test on data combined from all testing days.

We calculated a choice score to determine the magnitude of the difference in perceived naturalness of the tactile percepts evoked by biomimetic and linear ICMS encodings for a given electrode or electrode group across all test days. This score was calculated as:

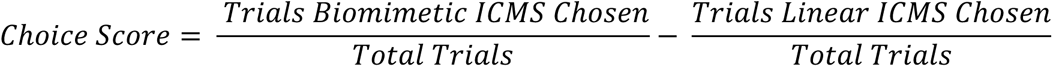

A choice score of 1 indicates that in all trials, the interval containing biomimetic ICMS was chosen, while a choice score of -1 indicates that the interval containing linear ICMS was chosen for every trial. A choice score of 0 indicates that an equal number of biomimetic and linear trials were selected. To determine if the distribution of choice scores for either simple or advanced ICMS differed significantly from 0, we used a Wilcoxon signed rank test. To determine if the distributions of choice scores for the simple and advanced ICMS trains differed from each other, we used a Wilcoxon rank sum test.

For the intensity matched linear trains, we used a Wilcoxon signed rank test to test whether the maximum amplitude and total charge were different than the equivalent amplitude and charge in the biomimetic ICMS trains. Lastly, a Friedman’s test was used test if there was any difference across days between the selected maximum linear amplitude in ICMS magnitude matching, as well as for the proportion of total trials that biomimetic ICMS was selected.

All data analysis and statistical tests were performed in MATLAB (MathWorks, Natick, MA). Normality was assessed with the Lilliefors test. P-values less than 0.05 were considered significant.

## 3. Results

### 3.1 Biomimetic ICMS encoding improves percept naturalness and uses less charge

Stimulus patterns that captured elements of the natural cortical response to mechanical indentations of the skin (figure 2(a) top), were chosen as feeling more like a mechanical stimulus than their linear stimulus counterparts. This was reflected by the positive distribution of choice scores across electrodes, with a median score of 0.18 (p = 0.022, Wilcoxon signed rank test, figure 2(b), dashed line). Across the 28 electrodes tested in three people, biomimetic ICMS felt more natural on 9 electrodes, while linear ICMS felt more natural on three electrodes. Importantly, every participant identified some electrodes for which biomimetic stimulation felt the most natural (figure 2(c-e) green bars). Only one participant (P2) had electrodes where simple linear ICMS felt like a better representation of the mechanical stimulus (figure 2(d), purple bars). To test whether these effects were reproducible, we repeated the experiment with the same electrodes across at least three days on most electrodes and found that in all these cases there was no difference in the proportion of trials in which the simple biomimetic stimulus was selected (P2, p = 0.80; P3, p = 0.95; C1, p = 0.63, Friedman’s test, figure 2(c-e) daily performance indicated by black circles). Overall, using a simple biomimetic ICMS train was a better perceptual representation of an actual mechanical stimulus for 32% of the electrodes, and there was no statistically significant difference for 57% of the electrodes. We examined the spatial distribution of the electrodes that evoked significant perceptual effects but found no obvious clustering or patterning for either electrode position (supplementary figures 1(a-c)) or projected field location (supplementary figure 2).

To determine if the ability of biomimetic ICMS trains to represent mechanical stimuli came at the cost of increased stimulation requirements (i.e. higher amplitude or total current delivered), we analyzed the pulse composition of the intensity matched trains. We found that the maximum amplitude (the highest amplitude pulse delivered during a train) was lower with the linear ICMS for two participants (P3, 66 μA, p < 0.001; C1, 76 μA, p = 0.0014; Wilcoxon signed rank test), but that there was no difference when considering all participants together (p = 0.69, Wilcoxon signed rank test, figure 3(a)). Additionally, when comparing the total charge between intensity matched trains, we found that more charge was delivered during the linear trains in two of the three participants (P2, 3.6 μC, p < 0.00l; C1, 3.1 μC, p < 0.00l) as well as across the entire group (p < 0.001, Wilcoxon signed rank test, figure 3(b)) compared to the biomimetic train (2.8 μC). This increase in charge was caused by the large difference in the pulse amplitudes during the hold phase; during this phase the rate of charge increase is minimized in biomimetic trains but maximized in linear trains (figure 3(c)). Consequently, biomimetic ICMS results in more natural sensations that also require less charge than linear ICMS trains.

**Figure 3.**
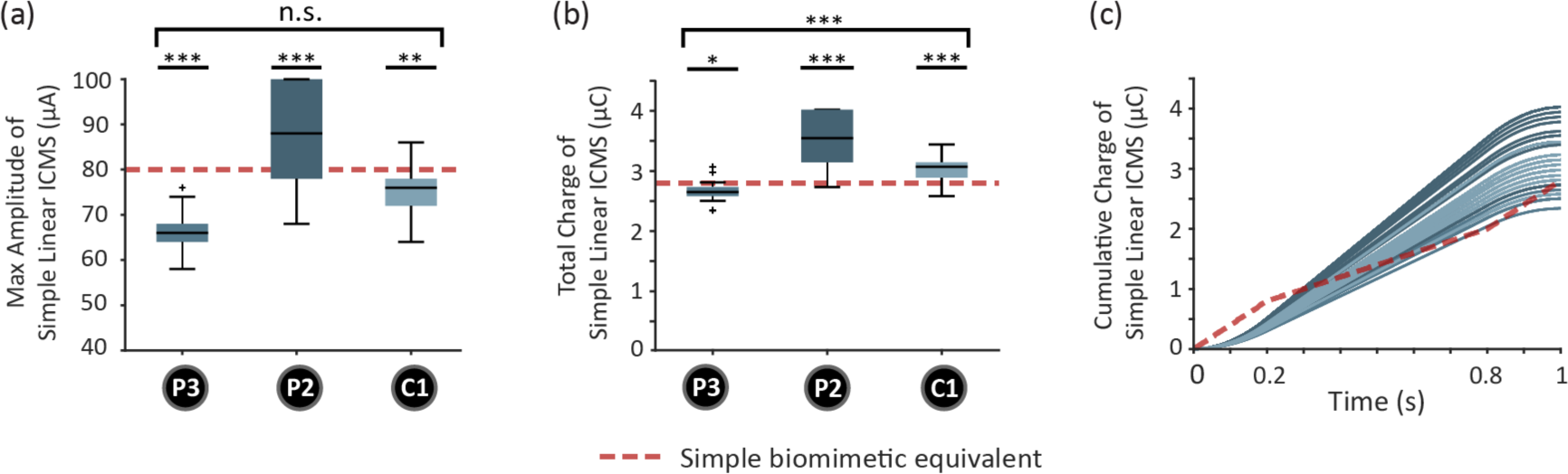
Effect of simple linear and biomimetic encoding on pulse amplitude and total charge. (a) Boxplots showing the distributions of stimulation amplitudes for simple linear trains that were selected by participants to match the perceived intensity of a simple biomimetic train. Data are combined from all testing days and electrodes for each participant. The red dashed line is set at the maximum amplitude used in the biomimetic trains. Outliers are shown with a cross. Participant identifiers are included in black circles. Statistical markers are shown for all participants combined, as well as for each individual participants. (b) Same as (a) but showing the total charge of simple linear ICMS train with a red dashed line corresponding to the total charge of the simple biomimetic ICMS train. (c) Cumulative charge of the simple linear ICMS trains over time, with the red dashed line representing the cumulative charge for the reference biomimetic ICMS train. The 0.2 s transient bursts in the biomimetic ICMS trains cause a rapid increase in cumulative charge, while simple linear ICMS had an initially low, but accelerating increase in charge that stabilized at a constant increase during the hold phase of the linear stimulation that contains high amplitude pulses. Statistical markers: ***p < 0.001, **p < 0.01, *p < 0.05, Wilcoxon signed rank test.

**Figure 4.**
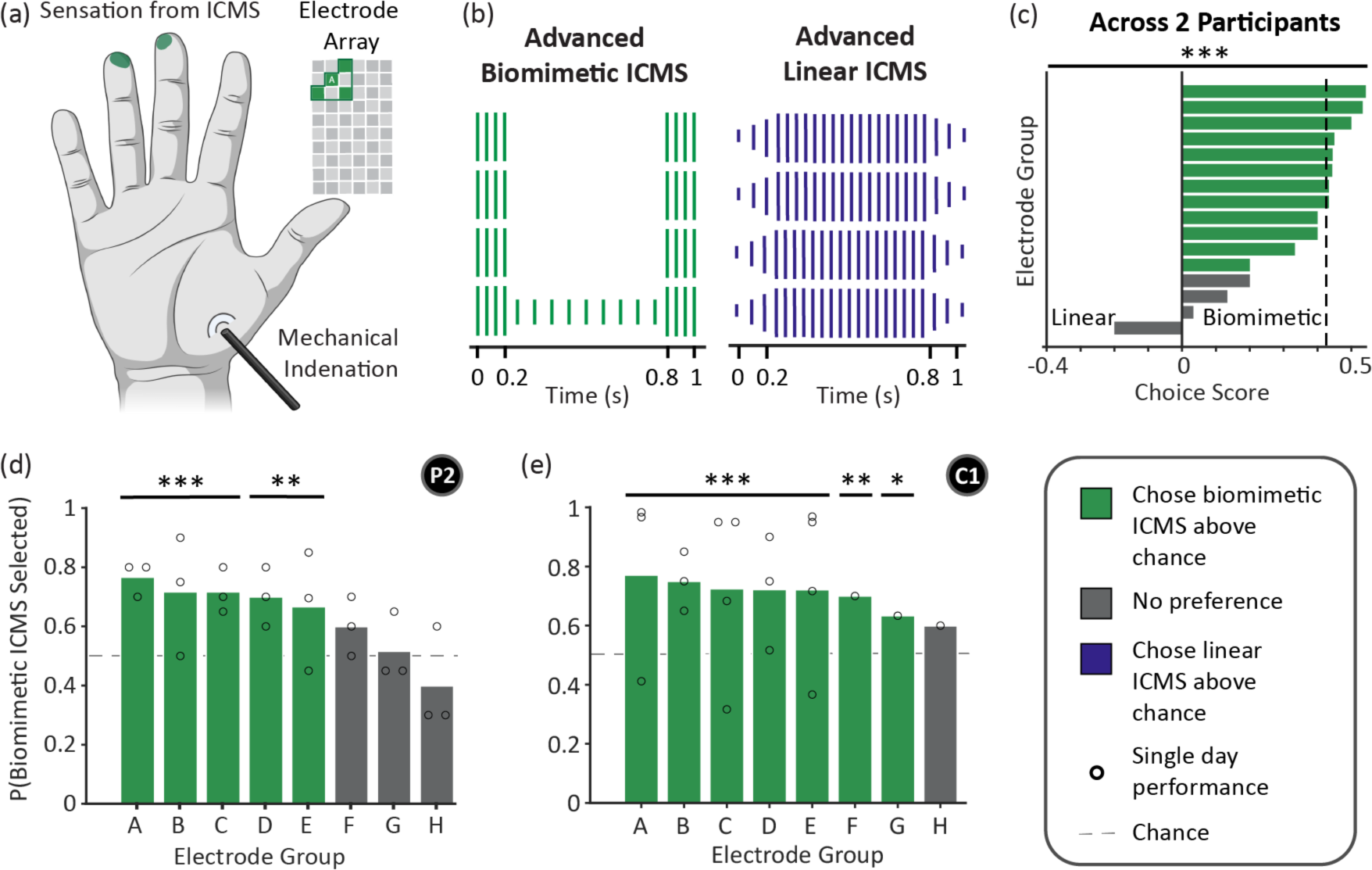
The effect of advanced biomimetic and linear ICMS on the naturalness of ICMS-evoked tactile percepts. (a) An example of advanced ICMS that was delivered across four electrodes that were adjacent on the electrode array. These four electrodes resulted in a tactile percept that spanned two adjacent regions of the fingertips. (b) Advanced ICMS encoding delivered stimulation using four electrodes while modulating both frequency and amplitude. Pulse frequency is represented by the spacing between the colored lines, while pulse amplitude is represented by the height of the colored lines. (Left) Advanced biomimetic ICMS had high-amplitude high-frequency bursts of stimulation during the 0.2 s indentation ramps, and stimulation during the hold-phase was delivered through just one electrode at a lower frequency and amplitude. (Right) The amplitude and frequency of advanced linear ICMS trains increased linearly with the increase in indentation depth. (c) Choice scores for each group of electrodes. Each bar indicates one group of electrodes. The dashed line indicates the median choice score value. Bars are colored as in Figure 2. Friedman’s test was used to determine if the overall choice scores differed from 0, indicating which ICMS encoding scheme produced more natural sensations. (d-e) Proportion of trials where biomimetic ICMS was selected across per electrode group. Bar coloring is the same as above. Black circles show the proportion of trials where biomimetic ICMS was selected on a single day (P2: n = 20 trials, C1: n = 60 trials, ***p < 0.001, **p < 0.01, *p < 0.05, Chi-squared goodness of fit test). Participant identifiers are indicated in black circles.

### 3.2 Encoding additional biomimetic features further improves percept naturalness

While simple biomimetic ICMS trains were better perceptual representations of natural mechanical stimuli than linear ICMS trains, the effect was not seen on all electrodes. We sought to test whether capturing more features of the natural cortical response to indentations might improve the effect of biomimetic ICMS. Specifically, contact transients during natural tactile stimuli evoke activity across broad areas of the somatosensory cortex as well as rapid changes in the firing rate of neurons [23]. Consequently, we extended the biomimetic approach to include multi-electrode stimulation that mimicked the spatiotemporal patterns of cortical responses and further modulated the stimulus frequency during the ramp and hold phases (figure 4(a)). Thus, in these advanced linear and biomimetic encoding schemes, we used both multi-electrode stimulation and frequency-amplitude co-modulation (figure 4(b)). We then repeated the previous task in which participants were asked to report which ICMS scheme felt most like the mechanical indentation.

Much like the prior result, we found that advanced biomimetic ICMS evoked tactile percepts that were a better perceptual match to the natural mechanical stimulus. However, unlike the simple ICMS schemes, the choice scores with advanced biomimetic ICMS were overwhelmingly positive; 75% of the electrode groups had significantly positive scores with a median score of 0.42 (p < 0.001, Wilcoxon signed rank test, figure 4(c), dashed line represents median). This increase in median choice scores from 0.18 for simple ICMS (figure 2(c)) to 0.42 for advanced ICMS (figure 4(c)) was significant (p = 0.0083, Wilcoxon rank sum test). For most of the individual electrode groups, advanced biomimetic ICMS was selected as being the better perceptual match to the mechanical stimulus well above chance (figure 4(d) and (e)). Additionally, in no cases did the advanced linear ICMS feel more natural. Supporting the robustness of this effect, the proportion of trials where advanced biomimetic ICMS was selected remained consistent across all testing days for both participants (P2, p = 0.79; C1, p = 0.76, Friedman’s test, figure 4(d) and (e), individual day performance indicated by black circles).

In participant C1, advanced ICMS evoked more intense percepts that required increases in the pre-indentation of the mechanical indenter so that the mechanical and electrical stimuli had similar intensities. Data were collected on four electrode groups prior to this change; after the change, there was a significant increase in the number of trials in which biomimetic ICMS was selected as a better match to the mechanical stimulus (p = 0.038, Friedman’s test, supplementary figure 4). Nevertheless, data from all days were included in our evaluation of whether linear or biomimetic stimulation trains were better perceptual representations of the mechanical indentation (figure 4(c,d,e)). The spatial distribution of the groups of electrodes on the array (supplementary figures 1(d) and (e)) or their projected field (supplementary figure 3) had no obvious relationship with the ICMS scheme that felt more natural.

We again asked whether the amplitude and charge were different between the linear and biomimetic ICMS trains for these advanced encoding schemes. The maximum amplitude of the intensity-matched linear ICMS trains were either no different or lower than the maximum amplitude of the biomimetic ICMS train (P2, 80 μA, p = 0.13; C1, 75 μA, p < 0.001; figure 5(a)). However, the advanced biomimetic ICMS trains required much less charge (5.5 μC) than the advanced linear trains (P2, 9.5 μC, p < 0.001; C1, 8.9 μC, p < 0.001; Wilcoxon signed rank test, figure 5(b)). The effect on charge was even more accentuated with these advanced trains as the rate of charge increase was dramatically lower during the hold phase for the biomimetic trains compared to the linear trains (figure 5(c)). Ultimately, advanced biomimetic ICMS evoked far more natural percepts while using substantially less charge.

**Figure 5.**
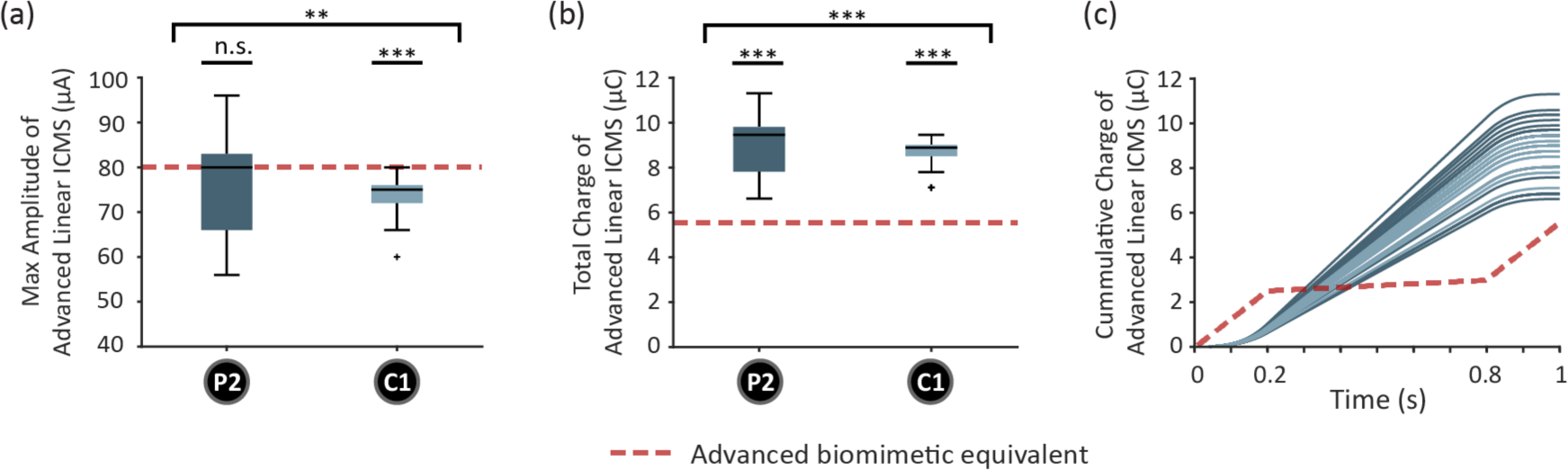
Effect of advanced linear and biomimetic encoding on pulse amplitude and total charge. (a) Boxplots showing distributions of stimulation amplitudes for advanced linear trains that were selected by participants to match the perceived intensity of an advanced biomimetic train. Data are combined from all testing days and electrodes for each participant. The red dashed is at the maximum amplitude used in biomimetic trains. Outliers are shown with a cross. Participant identifiers are included in black circles. (b) Same as (a) but showing the total charge of the advanced linear ICMS train. The red dashed line shows the total charge in the advanced biomimetic ICMS train. (c) Cumulative charge of the advanced linear ICMS trains over time, with the red dashed line representing the cumulative charge for the advanced biomimetic ICMS train. Statistics are shown for all participants combined, as well as for each individual participant. (***p < 0.001, **p < 0.01, *p < 0.05, Wilcoxon signed rank test).

### 3.3 Verbal descriptions of ICMS

The primary purpose of these experiments was to quantitively measure naturalness by having participants identify whether linear or biomimetic encoding schemes were better perceptual matches to a natural mechanical indentation. We did this to avoid relying solely on qualitative verbal reports. Nevertheless, the participants did sometimes describe what they felt during these experiments. Examples of the verbal reports included: “It felt more like the touch and then pressure of the indenter” – P3; “The sensation felt more focal like an actual poke, with a clear start and stop, and less warm” – P2; “Usually the double bump felt similar to the mechanical stim but not always.” – C1.

## 4. Discussion

### 4.1 Biomimetic encoding increases the naturalness of ICMS-evoked sensations

A major challenge for electrical stimulation systems is the very large parameter space. Even in simple systems, pulse width, pulse amplitude, and frequency can be modified, all of which have measurable and distinct consequences. Indeed, pulse frequency alone has effects on the conscious percepts evoked by ICMS, which can vary from electrode to electrode [19]. Therefore, if specific pulse timing is important [36], if spatiotemporal dynamics matter, and as the number of stimulation electrodes increases, the total parameter space rapidly becomes intractable. A conceptual framework is required to design general purpose stimulation patterns. Here, we chose to use a biomimetic framework to ask whether stimulation trains were better perceptual matches to an actual mechanical stimulus than stimulation trains that simply reflected the mechanics of the input itself. The rationale for using a biomimetic encoding scheme is the intuition that artificial activation of the cortex that reproduces the most salient patterns of cortical neural activity during natural touch will activate populations of cortical neurons in a way that more closely mimics their normal responses, and thus evoke intrinsically more natural percepts [11,37,38].

There is precedent for this from both modelling and experimental studies. In simulated cortical columns, biomimetic stimulation evokes patterns of neural activity that exhibit characteristics of natural neural responses [39]. On the experimental side, calcium imaging in rodents has shown that biomimetic ICMS pulse trains can produce more natural patterns of cortical activity, suggesting that efforts in humans could produce similar results [22]. Further, biomimetic ICMS in humans improves the resolution of the tactile signal and mitigates some of the decoding challenges caused by stimulation [15]. In the peripheral nervous system, biomimetic stimulation can improve both the perceptual naturalness of artificial electrical stimulation [10] and improve functional performance of a prosthesis [12].

Here, we endeavored to incorporate both the rapid increases in the volume of cortical tissue activation and neural spiking rate that are associated with the onsets and offsets of mechanical indentations [23]. Our results with both simple and advanced biomimetic ICMS are consistent with the hypothesis that artificially evoked neural activity that resembles naturally evoked neural activity is more likely to be appropriately interpreted by the brain. Additionally, the fact that our findings are consistent across participants, and did not require any learning, implies that this is a robust and ubiquitous principle. Furthermore, our results were consistent across different locations on the array and somatotopic regions of the hand (palm and digit tips), indicating the robust benefits of biomimetic stimulation (supplementary figures 1-3). Ultimately, these results demonstrate that using naturally evoked patterns of neural activity is a useful template for designing biomimetic stimulation encoding schemes to recreate percepts that we feel in our everyday lives. Further, this framework could substantially reduce the effort associated with manually exploring the large stimulus parameter space.

### 4.2 ICMS-evoked sensation naturalness increases with the level of biomimicry

In the first set of experiments, we chose to focus on a single dimension of biomimicry; the spatial changes in cortical activation that occur during the onset and offset phases of a ramp-and-hold stimulus [23]. We used amplitude modulation through a single electrode to drive changes in the area of cortical activation. Further, we simplified the problem even more and used a single stimulus amplitude to represent the two transient phases, and a lower amplitude to represent the hold phase. A natural question that emerged was whether increasingly biomimetic stimulation patterns would result in increasingly natural perceived sensations. Our findings imply that the degree of biomimicry is in fact related to the perceived naturalness. Indeed, we observed that the proportion of electrodes or electrode groups that evoked more natural percepts with biomimetic ICMS increased from 32% (figure 2(c)) to 75% (figure 4(c)) when shifting from a simple to a more advanced encoding scheme that captured more features of the cortical response of touch. Furthermore, this change from a simple to advanced encoding scheme increased the magnitude of the preference for biomimetic ICMS, as reflected in the increase in choice scores (figure 2(c) and 4(c)).

Our experimental design did not include a direct comparison between simple and advanced ICMS due to the difficulty of intensity matching between single and multi-electrode stimulation. The relative importance of individual features (such as multi-electrode versus co-modulation of amplitude and frequency) remains untested, as does the contribution of overall sensation intensity to the perception of naturalness. Based upon work in rodents, it is possible that ICMS with frequency modulation alone would not produce a strong effect on perceived naturalness as it does not induce transient bursts in neural activity [22]. Additionally, the participant C1 reported that qualitative comparisons between the ICMS and mechanical stimulus was difficult when the intensities of the two were substantially different, something that motivated our additional intensity matching efforts (supplemental figure 4). Future experiments performing more systematic comparisons of stimulation parameters might reveal the relative importance of these features and inform future stimulation design. In the peripheral nervous system, incorporating additional responses of the neural activation from different types of peripheral mechanoreceptors can lead to changes in the perceived naturalness of the artificial stimuli [10].

### 4.3 Biomimetic ICMS may extend array health

A key consideration with electrical stimulation of the nervous system is safety and device longevity [40]. For ICMS, stimulation charge has a major impact on stimulation-induced neuronal damage [41,42]. We discovered that both simple and advanced biomimetic ICMS were able to evoke sensations of equal intensity with less charge (figure 3 and 5). This finding held true across multiple participants, electrodes, and days. Biomimetic ICMS may therefore improve the longevity of implanted devices due to increasing charge efficiency, thus reducing power consumption while also enhancing naturalness, leading to improved user experience, and increased practicality for long-term use for sensory feedback in neuroprosthetics.

### 4.4 Implications for bidirectional neuroprosthetics

In this study, we focused only on recreating neural patterns from a simple, predefined mechanical indentation to the palm. Ideally, we would be able to recreate a variety of different perceived sensations that we feel every day, from vibrations associated with different textures to stable percepts during extended contact [37]. Having found that representing the natural neural activity improves naturalness for a simple peripheral input, it is possible that using this biomimetic framework will allow us to develop more complicated stimulation patterns from a wider range of tactile experiences to make more informed biomimetic encoding algorithms.

While it has been shown that biomimetic ICMS has multiple functional benefits, from improving discriminable force levels to minimizing motor decoding with lower levels of sustained stimulation [15,24], it remains unclear how more natural percepts from ICMS may impact performance of bidirectional brain-computer interfaces. Previous evidence from peripheral nerve stimulation indicates that not only does biomimetic ICMS increase perceived naturalness, but it also improves resiliency to cognitive distractors during motor tasks and improves embodiment of neuroprosthetics [10,11,43]. Future experiments will show if the improved naturalness we achieved in this study will have similar functional benefits for bidirectional brain-computer interfaces.

### 4.5 Limitations

There were several limitations in this study. First, we were unable to deliver mechanical indentations to the exact location where ICMS evoked percepts in all our participants. This was due to combinations of SCI-induced sensory loss and hand rigidity, which made delivering mechanical stimulation to the digits impossible. The locational difference between the mechanical reference and the artificial sensation could have complicated the task, but linear and biomimetic ICMS trains were subject to this same potential challenge. If possible, future studies should explore if increasing the ease of comparison affects the results we found here.

Second, each of the participants has varying degrees of reduced sensation as a result of their SCI. While each of the participants had monofilament thresholds within the natural range on their thenar eminence during their initial clinical evaluation, there may be indirect effects on their perception of the stimuli. However, even if each participant had uniquely altered perception of the stimuli, this would not explain why they unanimously preferred the biomimetic encoding.

A third limitation, which is a conceptual challenge for biomimetic electrical microstimulation more generally, is that with existing technology, it is impossible to activate neurons in a way that directly reflects what might be recorded from a microelectrode. Currently, neurons around the electrode tip are all activated in synchrony with each ICMS pulse [31]. So, while we can use biomimetic ICMS to improve naturalness, we are unable to reproduce percepts that might depend on detailed patterns of neural activity that occur within small regions of the cortex [31,32,44]. Even with this limitation, our focus is not driven by perfectly replicating perceived sensations, but to drive meaningful improvements in day-to-day life for the millions that would benefit from neuroprosthetics.

## 5. Conclusion

We investigated how incorporating the key elements of activity in the somatosensory cortex in response to indentations on the hand improved the naturalness of ICMS-evoked percepts. We directly measured naturalness by having three human participants compare the artificial percepts from ICMS to a naturally occurring sensation on their hand. We found that even at the single electrode level, there was a tendency for the participants to prefer biomimetic ICMS trains and that this preference was enhanced as additional features of the natural cortical response to mechanical indentations were included in the stimulus train design. These findings support the notion that biomimetic stimulation is a key framework to improving artificial tactile feedback for neuroprosthetics.

## Acknowledgements

We would like to acknowledge our participants for their exceptional effort, commitment, and insightful discussions; Debbie Harrington (Physical Medicine and Rehabilitation) for the regulatory management of this study; Gracie Hilber and Carleigh May for their help with data acquisition; Willam Hockeimer for his advisement on statistical analysis. We would also like to express our heartfelt gratitude to Prof. Bensmaia, who sadly passed away during this study. His unwavering support, insightful guidance, and immense passion for natural touch and biomimicry were instrumental to our work. This paper is dedicated to his memory, honoring his remarkable contributions to science and his lasting impact on our team and the broader scientific community. This work was supported by the National Institutes of Health (NIH) through National Institute of Neurological Disorders and Stroke grants UH3 NS107714 and R35 NS122333.

## Data availability statement

The data supporting this study’s findings are available upon reasonable request from the authors.

---

**Supplementary Figure 1.**
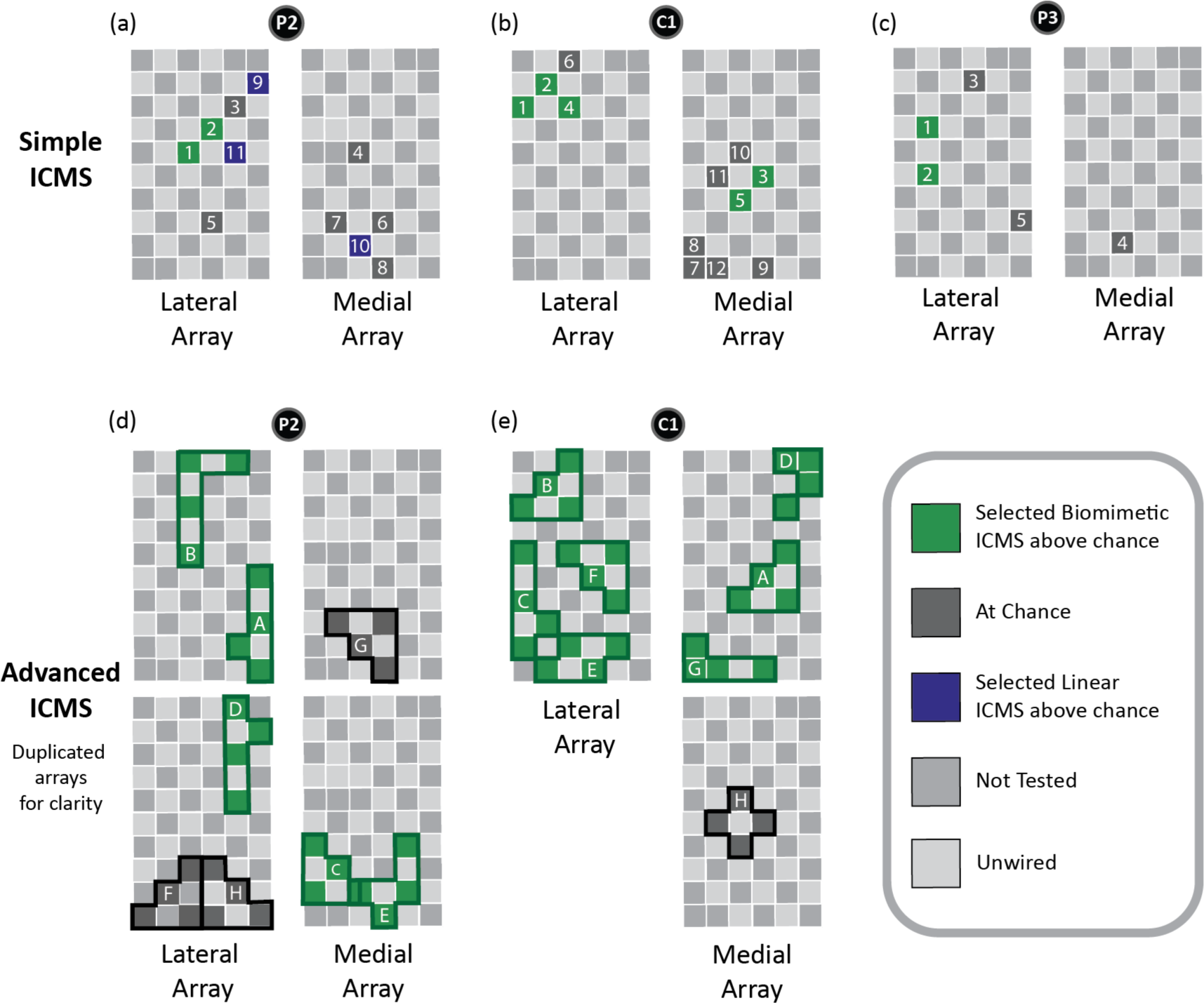
Electrode location was not related to which ICMS encoding scheme was chosen as feeling more like the mechanical reference. (a-c) Location of tested electrodes on the two arrays for each participant. Each electrode is colored based on whether one of the stimulus encoding schemes was selected as a better perceptual representation of the mechanical stimulus above chance, combined across all testing days (green = biomimetic trials above chance, gray = at chance, purple = linear trials above chance, p < 0.05, Chi-square goodness-of-fit test). (d-e) Location of groups of electrodes on the two arrays for P2 and C1. Arrays are duplicated in a column due to some electrode overlap between stimulation groups. The group letter is located on the electrode that maintained constant stimulation for the entire second. Electrodes are colored as in (a-c). Participant identifiers included in black circles.

**Supplementary Figure 2.**
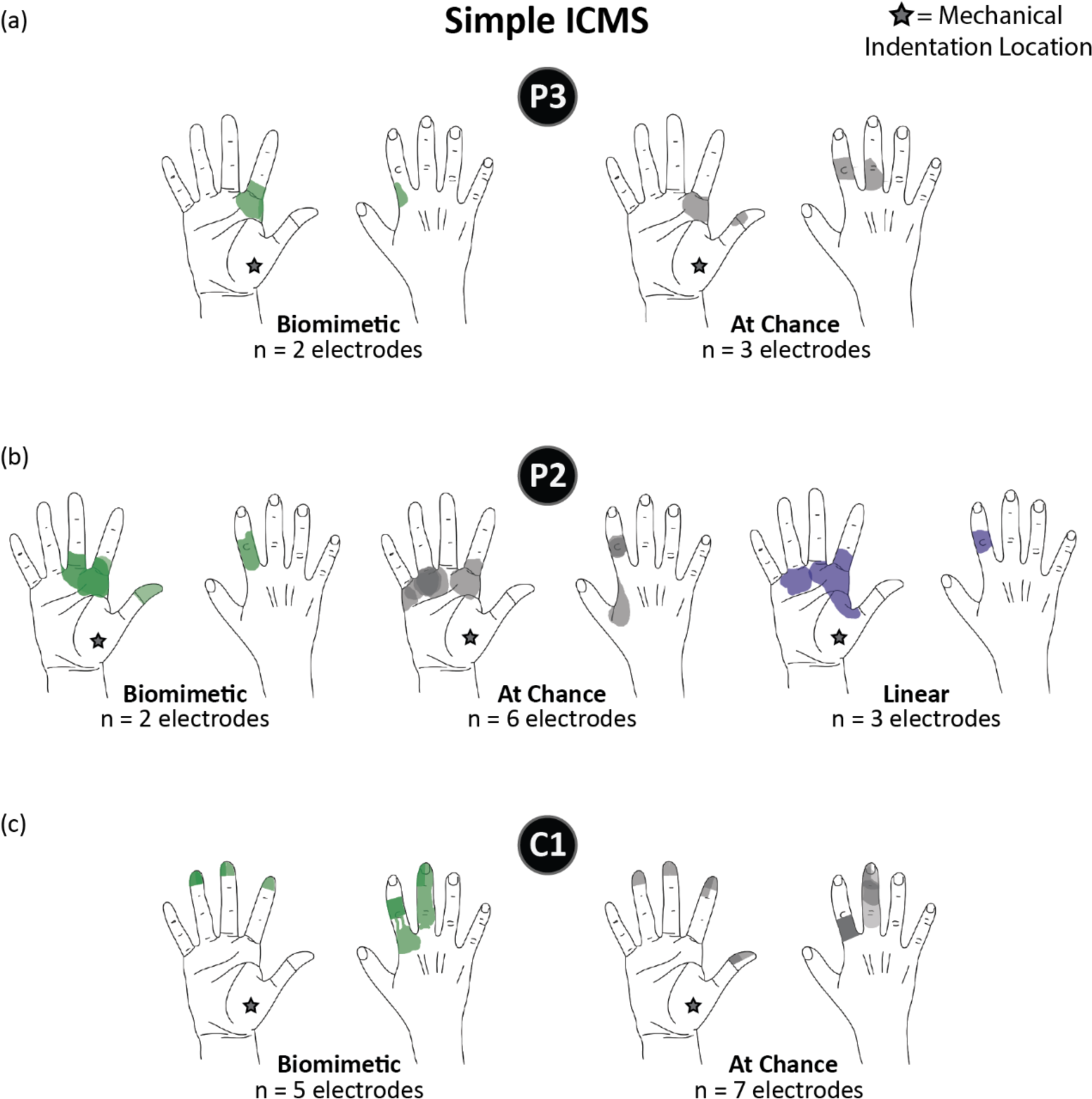
Projected field overlap between electrodes that evoked different perceptual choices for simple ICMS encoding schemes. (a-c) Colored regions indicate the location on the hand where 60 μA, 100 Hz, 1 s ICMS trains were perceived by each participant. Each color indicates the ICMS encoding scheme that was chosen to best match the mechanical stimulus (green = biomimetic, gray = at chance, purple = linear, Chi-square goodness-of-fit test). The star symbol indicates the approximate location of the mechanical indentation. Participant identifiers are included in black circles.

**Supplementary Figure 3.**
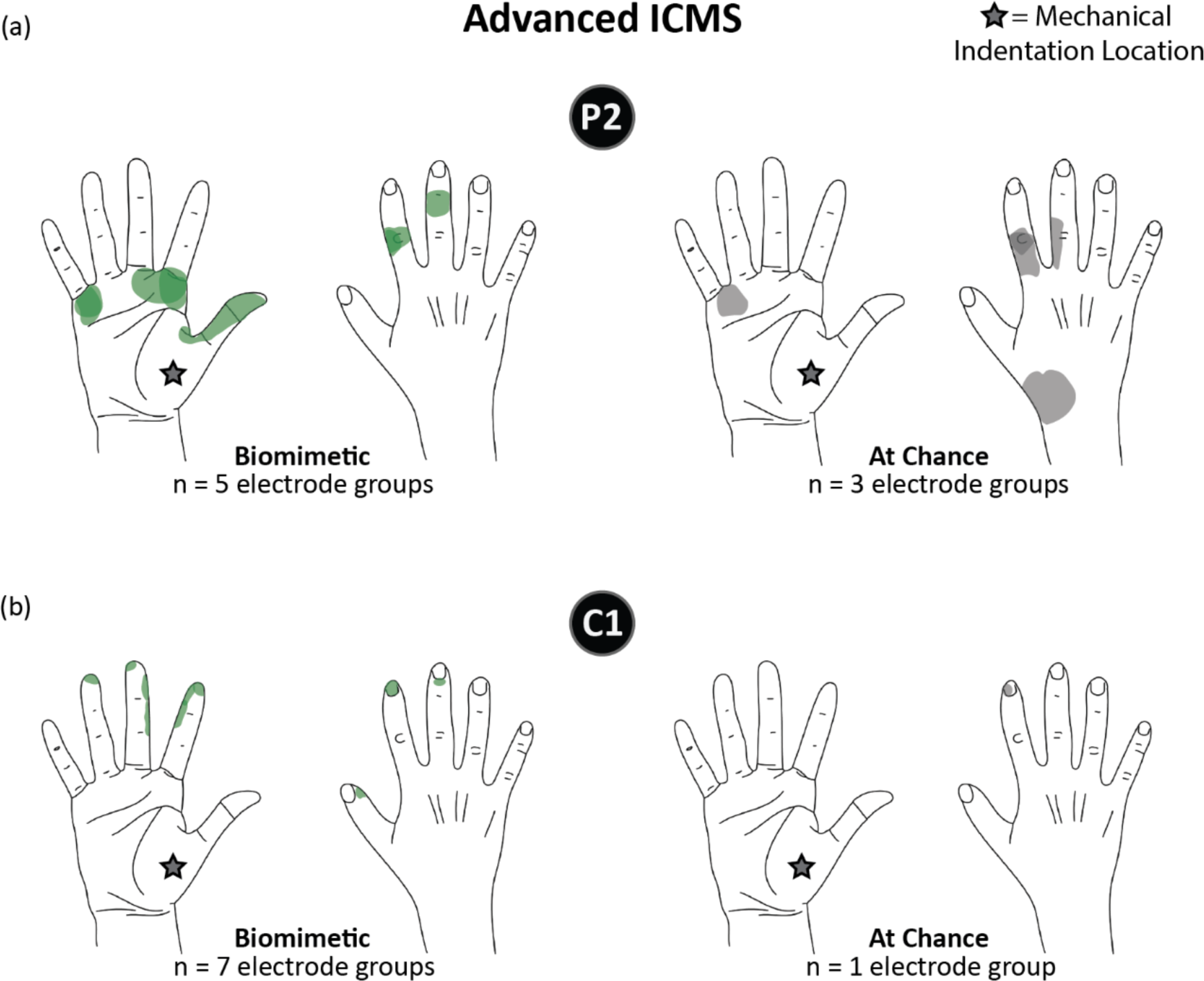
Projected field overlap between electrode groups that evoked different perceptual choices for advanced ICMS encoding schemes. (a-c). Colored regions indicate the location on the hand where 60 μA, 100 Hz, 1 s ICMS trains across a group of four electrodes were perceived by each participant. Each color indicates the ICMS encoding scheme that was chosen to best match the mechanical stimulus (green = biomimetic, gray = at chance, purple = linear, Chi-square goodness-of-fit test). The star symbol indicates the approximate location of the mechanical indentation. Participant identifiers are included in black circles.

**Supplementary Figure 4.**
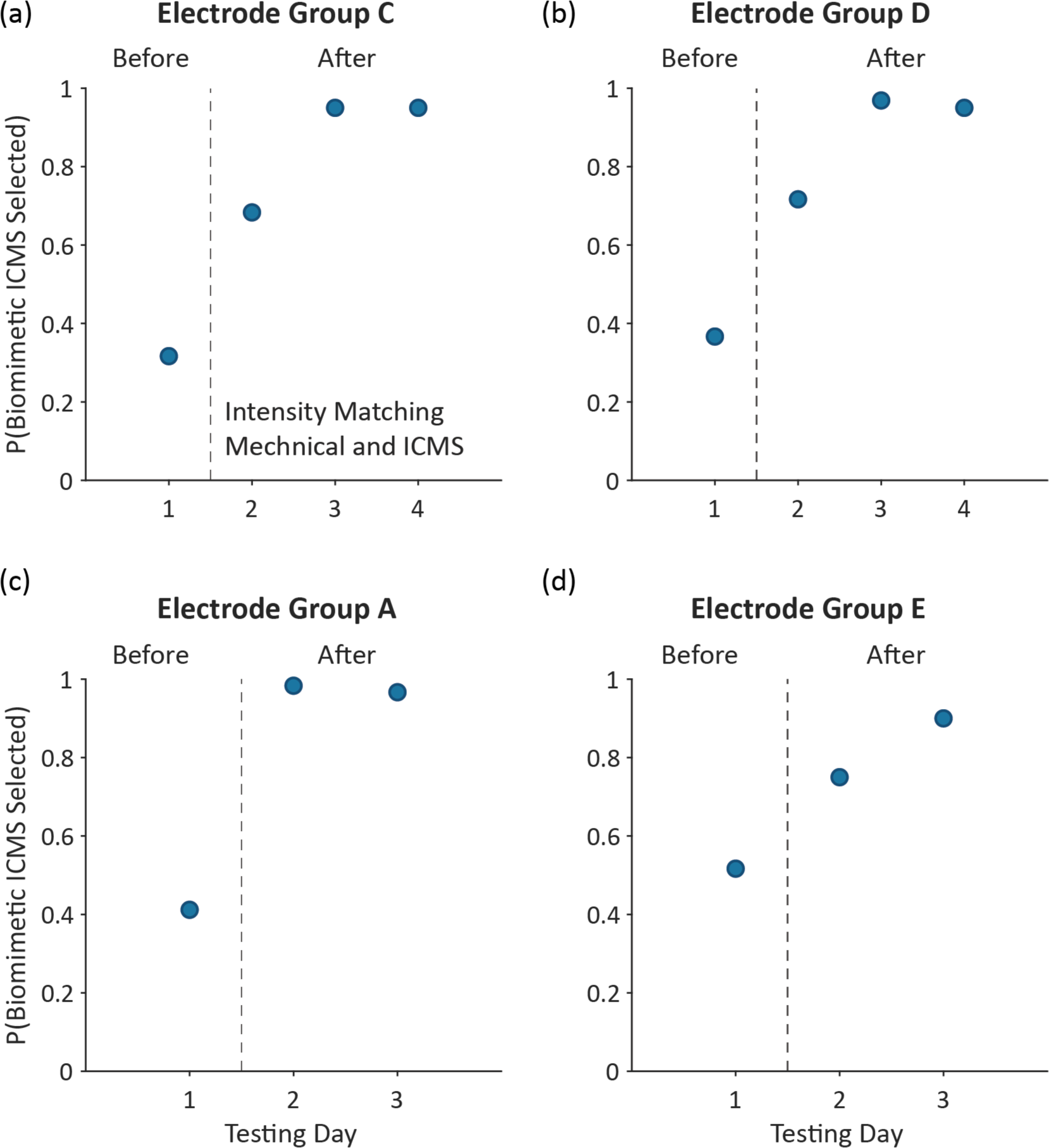
Effect of intensity matching method on performance for advanced ICMS in participant C1. (a-d) The proportion of trials where advanced biomimetic ICMS was selected across testing days for four electrode groups in participant C1. The dashed line indicates when intensity matching between mechanical and ICMS was improved.

